# Machine learning-based mortality prediction models for non-alcoholic fatty liver disease in the general United States population

**DOI:** 10.1101/2024.07.10.24310253

**Authors:** Jia-Rui Zheng, Zi-Long Wang, Bo Feng

## Abstract

**Background & Aims:** Nowadays, the global prevalence of non-alcoholic fatty liver disease (NAFLD) has reached about 25%, which is the most common chronic liver disease worldwide, and the mortality risk of NAFLD patients is higher. Our research created five machine learning (ML) models for predicting overall mortality in ultrasound-proven NAFLD patients and compared their performance with conventional non-invasive scoring systems, aiming to find a generalizable and valuable model for early mortality prediction in NAFLD patients.

**Methods:** National Health and Nutrition Examination Survey (NHANES)-III from 1988 to 1994 and NHANES-III related mortality data from 2019 were used. 70% of subjects were separated into the training set (N = 2262) for development, while 30% were in the testing set (N= 971) for validation. The outcome was all-cause death at the end of follow-up. Twenty-nine related variables were trained as predictor features for five ML–based models: Logistic regression (LR), K-nearest neighbors (KNN), Gradient-boosted decision tree (XGBoost), Random forest (RF) and Decision tree. Five typical evaluation indexes including area under the curve (AUC), F1 score, accuracy, sensitivity and specificity were used to measure the prediction performance.

**Results:** 3233 patients with NAFLD in total were eligible for the inclusion criteria, with 1231 death during the average 25.3 years follow up time. AUC of the LR model in predicting the mortality of NAFLD was 0.888 (95% confidence interval [CI] 0.867-0.909), the accuracy was 0.808, the sensitivity was 0.819, the specificity was 0.802, and the F1 score was 0.765, which showed the best performance compared with other models (AUC were: RF, 0.876 [95%CI 0.852-0.897]; XGBoost, 0.875 [95%CI 0.853-0.898]; Decision tree, 0.793 [95%CI 0.766-0.819] and KNN, 0.787 [95%CI 0.759-0.816]) and conventional clinical scores (AUC were: Fibrosis-4 Score (FIB-4), 0.793 [95%CI 0.777-0.809]; NAFLD fibrosis score (NFS), 0.770 [95%CI 0.753-0.787] and aspartate aminotransferase-to-platelet ratio index (APRI), 0.522 [95%CI 0.502-0.543]).

**Conclusions:** ML–based models, especially LR model, had better discrimination performance in predicting all-cause mortality in patients with NAFLD compared to the conventional non-invasive scores, and an interpretable model like Decision tree, which only used three predictors: age, systolic pressure and glycated hemoglobin, is simple to use in clinical practice.

## Introduction

NAFLD has become the most common chronic liver disease and affects up to 1 billion people worldwide, leading to a health and economic burden^1–3^, and can increase the risk of end-stage liver disease and hepatocellular carcinoma (HCC)^4^. Patients with NAFLD have a significant increase of the all-cause mortality, among which the main causes are cardiovascular disease, malignant tumor as well as end-stage liver disease^5, 6^. It is of great importance to early detect patients with a higher risk of death, which may may help to optimize the use of finite resources and provide appropriate care.

In addition to age, fibrosis stage has the best predictive power for overall mortality^7, 8^. However, liver biopsy as the gold standard is inappropriate to screen clinically significant fibrosis because of its features like invasive, inconvenient and expensive. Some studies have shown that conventional non-invasive scores, such as NFS^9^, FIB-4^10^, and APRI^11^, have prognostic significance of death for NAFLD patients^12–14^. However, their results were controversial. A meta-analysis including 19 longitudinal studies showed that only the NFS > 0.676 was predictive of overall mortality, while FIB-4 and APRI failed^14^. And a retrospective analysis including 646 NAFLD patients proven by liver biopsy revealed that although FIB-4 and NFS could precisely predict the risk of overall mortality of NAFLD patients, owing to the AUC values were not high enough (FIB-4, 0.72 [95% CI, 0.68–0.76]; NFS, 0.72 [95% CI, 0.68–0.76] and APRI, 0.52 [95% CI, 0.47–0.57]), so they were not useful in the clinical practice and new methods are needed to confirm the prognosis of NAFLD patients ^13^.

Predictive tools using ML have been extensively developed and used in medicine in recent years because they are often superior to traditional predictive methods^16^, and nowadays, the utilization of ML in gastroenterology territory is in steady-state growth^17^. Some studies have shown that ML is superior to traditional non-invasive approaches for the prediction of liver fibrosis^18–21^, such as FIB-4 and FibroScan to predict significant fibrosis (≥F2) and advanced fibrosis (≥F3) in NAFLD patients^18^. However, a model for predicting NAFLD mortality based on ML has not yet been developed. Our study aimed to develop, test and verify the mortality prediction model established by ML for patients with NAFLD in the USA.

## Methods

### Data sources and ethical approval

NHANES-III (1988–1994) database with nationwide, multilevel, stratified, clustered probability sampling design, is used to assess the health status of the civilian population in the USA. The data in the NHANES-III includes interviews, physical examinations, laboratory tests, and ultrasound examinations were conducted to assess the liver steatosis. NHANES-III data is also related to death certificates from the National Death Index (NDI) as of December 31, 2019, allowing for mortality analysis.

The survey was ratified by the ethics review committee of the National Center for health statistics, and the written informed consent of all participants was acquired to collect data. The institutional review committee dispensed with the consideration of human research because the data was fully certain.

### Study population and definitions

In the NHANES-III survey, among the adult participants (20-74 years old) with gradable liver / gallbladder ultrasound results (n=13,856), we first excluded individuals with heavy drinking (men >21drinks / week, women >14 drinks/ week), viral hepatitis (serum hepatitis B surface antigen positive and /or serum hepatitis C antibody positive), iron overload (transferring saturation≥50%). In addition, participants with incomplete or missing data on mortality, physical examinations and laboratory tests were also excluded (**Figure 1**).

**Figure 1.**
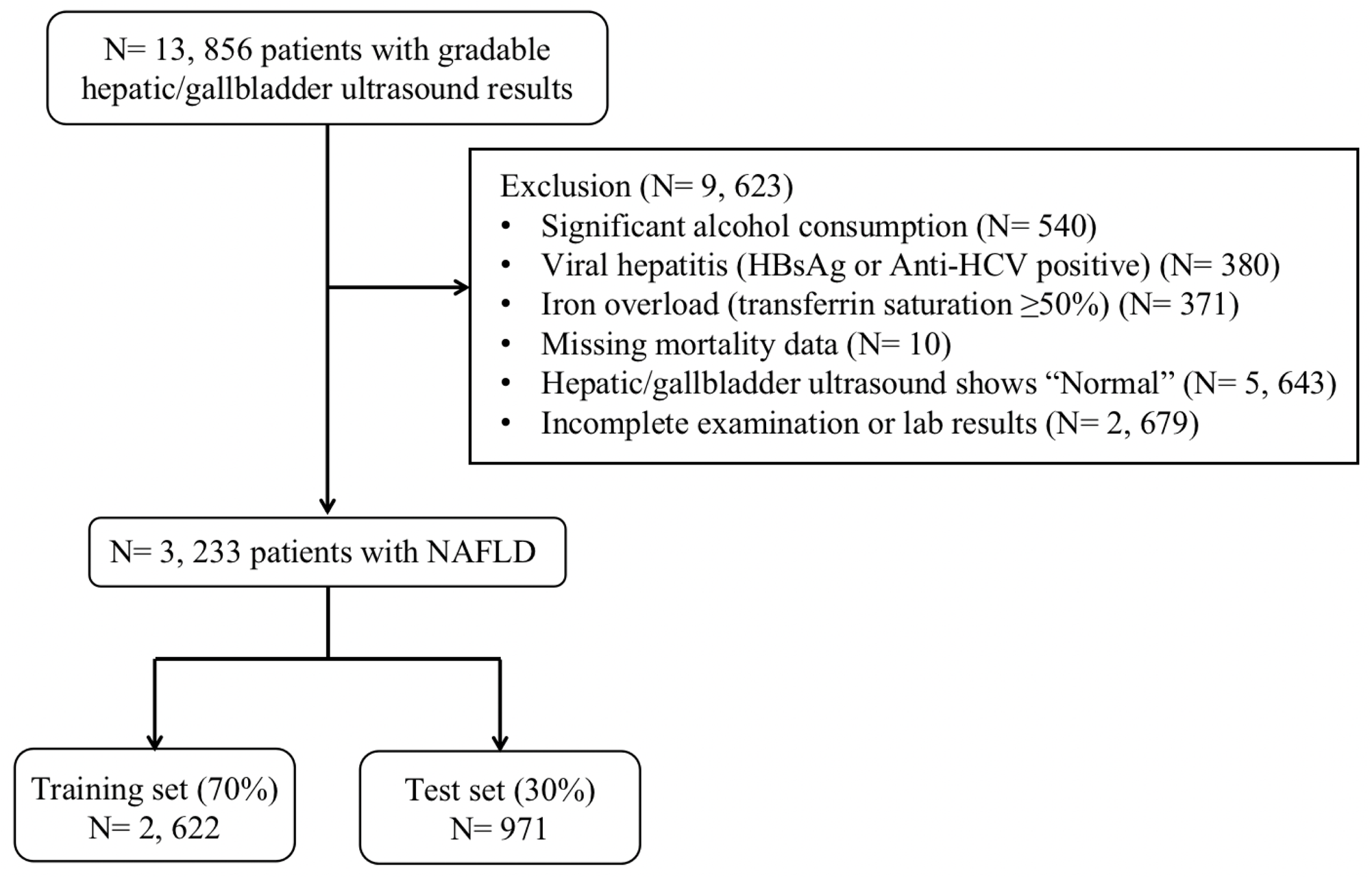
Study design and data partitioning flow chart. NAFLD, non-alcoholic fatty liver disease.

NHANES-III examination includes gallbladder ultrasonography in adults aged 20-74. In order to evaluate fatty liver, the gallbladder ultrasound images were examined by three committee certified radiologists. The following five criteria were used in the review process: (I) parenchymal brightness, (II) liver to kidney contrast, (III) deep beam attenuation, (IV) bright vascular walls, and (V) gallbladder wall definition. The degree of hepatic steatosis were reported as normal, mild, moderate or severe according to these five criteria. In this study, NAFLD was defined as mild to severe hepatic steatosis, excluding whatever known causes of liver disease.

### Variable selection and outcome

In this study, twenty-nine NAFLD related factors were included, such as demographic features (age, gender and ethnicity), general measurement (waist circumference, body mass index (BMI), systolic blood pressure (SBP) and diastolic blood pressure (DBP)), biochemistry tests (WBC, PLT, C-reactive protein (CRP), iron, total iron-binding capacity (TBIL), ferritin, transferrin saturation, total cholesterol, triglyceride, high-density lipoprotein (HDL) cholesterol, and uric acid), diabetes testing profile (fasting plasma glucose, glycated hemoglobin (HbA1c), fasting C-peptide and fasting insulin),and liver chemistry (aspartate aminotransferase (AST), alanine aminotransferase (ALT), alkaline phosphatase (ALP), gamma glutamyl transferase (GGT), albumin and total bilirubin).

The outcome was set as the passive mortality as of December 31, 2019 according to the follow up time of NHANES-III. For assessing the state of death (including the date of death), we performed probability matching with the NDI records. NHANES-III related death documents use the ucod_113 to code the deaths before 1998 and between 1999 and 2015, which are coded according to the Ninth Revision of the international classification of diseases (ICD-9).

### Development and validation of machine learning models

The ML models in our study, including LR, KNN, Decision tree, RF and XGBoost, were trained using the selected 29 variables to predict mortality, and then 10-fold stratified cross-validation was adopted in the training process to avoid overfitting of the mode. Briefly speaking, the training data was divided into 10 hierarchical subsets, followed by using 9 subsets to train the model, and using the left one subset for verification. These training and verification processes were repeated 10 times, and each subset was used once as the verification dataset, so that we could obtain 10 estimates of prediction accuracy, and these estimates were averaged to obtain a single estimate. For the LR model, we described the absolute value of standardized beta coefficient, while for the RF and XGBoost, the feature importance was showed of each model.

Testing data were used to verify the performance of developed ML models, which was independent from the training process. Accuracy, sensitivity or recall, specificity, precision, AUC and the F1 score (the harmonic average of recall and precision) were taken as performance indicators, and then compared with those of three conventional NAFLD scores (FIB-4, NFS, APRI) on the testing dataset, where we also established calibration plots in order to observe the coherence between predicted and observed mortality during follow-up. A good calibration degree shows that from the model explanation to the random samples prediction, the predicted value of the model is closer to the actual probability of the results.

### Statistical analysis

To present the patient characteristics, the mean of standard deviation (SD) for numerical variables and percentage counting for the categorical variables were used. We use Student t-test to compare the mean value between two samples, and chi-square test to compare the frequencies. For all tests, the bilateral significance level <5% was considered statistically significant (p<0.05). All statistical analyses were conducted using RStudio software (Macintosh; Intel Mac OS X 12_5_0).

## Results

### Characteristics of study subjects

3,233 patients with NAFLD met the inclusion criteria and were categorized into two groups at random: training set (70%, N = 2262) as well as testing set (30%, N = 971). **Figure 1** shows the patient screening process. The overall mortality in patients with NAFLD is 38.7% during the average 25.3 years follow up time. The baseline characteristics of patients according to whether died or not in the training and testing sets are described in **Table 1**. Compared with the patients finally died, those survival NAFLD patients were more likely to be young people, women, Mexican Americans. In addition, BMI, waist circumference, SBP, DBP, TC, TG, CRP, uric acid, GGT, ALP, HbA1c, FPG, fasting C-peptide, fasting insulin of these people were lower, and PLT, TIBC, ALT, albumin were higher. There were no significant differences between the training set and testing set for all factors.

**Table 1.** Baseline characteristics of the data set.

|  | Trainning data set (N=2262) |  |  | Testing data set (N=971) |  |  |
| --- | --- | --- | --- | --- | --- | --- |
|  | Nonsurvival (n=861) | Survival (n=1401) | P | Nonsurvival (n=370) | Survival (n=601) | P |
| **Demographic** |  |  |  |  |  |  |
| Age (yr) | 59.3 (12.2) | 38.8 (12.2) | <0.001 | 58.9 (12.9) | 37.9 (12.2) | <0.001 |
| Gender (male) (%) | 438 (50.9%) | 588 (42.0%) | <0.001 | 196 (53.0%) | 269 (44.8%) | 0.015 |
| Ethnicity |  |  | <0.001 |  |  | <0.001 |
| Mexican American (%) | 213 (24.7%) | 548 (39.1%) |  | 96 (25.9%) | 238 (39.6%) |  |
| Non-hispanic balck (%) | 222 (25.8%) | 350 (25.0%) |  | 88 (23.8%) | 153 (25.5%) |  |
| Non-hispanic white (%) | 396 (46.0%) | 441 (31.5%) |  | 173 (46.8%) | 173 (28.8%) |  |
| Others | 30 (3.48%) | 62 (4.43%) |  | 13 (3.51%) | 37 (6.16%) |  |
| **Body measurement** |  |  |  |  |  |  |
| Body mass index (kg/m2) | 30.4 (6.50) | 29.1 (6.63) | <0.001 | 30.3 (6.54) | 29.0 (6.83) | 0.002 |
| Waist circumference (cm) | 104 (14.3) | 95.9 (16.1) | <0.001 | 104 (14.3) | 96.0 (16.8) | <0.001 |
| Systolic blood pressure (mmHg) | 137 (19.6) | 120 (15.0) | <0.001 | 138 (20.3) | 120 (14.8) | <0.001 |
| Diastolic blood pressure (mmHg) | 78.1 (10.7) | 75.0 (10.4) | <0.001 | 77.5 (10.7) | 75.5 (10.3) | 0.003 |
| **Biochemistry tests** |  |  |  |  |  |  |
| WBC (×10<sup>9</sup>/L) | 7.62 (2.61) | 7.23 (1.98) | <0.001 | 7.36 (1.93) | 7.34 (2.12) | 0.860 |
| PLT (×10<sup>9</sup>/L) | 267 (71.0) | 279 (67.1) | <0.001 | 268 (69.7) | 285 (74.6) | <0.001 |
| Iron (ug/dL) | 14.5 (5.30) | 15.1 (6.02) | 0.021 | 15.0 (5.74) | 14.8 (5.72) | 0.702 |
| Total iron-binding capacity (ug/dL) | 63.2 (9.74) | 66.2 (10.4) | <0.001 | 63.4 (9.83) | 66.6 (10.3) | <0.001 |
| Transferrin saturation (%) | 23.5 (9.00) | 23.3 (9.57) | 0.780 | 24.0 (9.28) | 22.9 (9.46) | 0.082 |
| Ferritin (ng/mL) | 187 (340) | 129 (138) | <0.001 | 172 (167) | 125 (131) | <0.001 |
| Total cholesterol (mg/dL) | 5.75 (1.13) | 5.21 (1.11) | <0.001 | 5.55 (1.05) | 5.21 (1.09) | <0.001 |
| Triglyceride (mg/dL) | 2.26 (1.47) | 1.83 (1.42) | <0.001 | 2.12 (1.33) | 1.85 (1.37) | 0.002 |
| HDL cholesterol (mg/dL) | 1.21 (0.43) | 1.23 (0.37) | 0.276 | 1.21 (0.43) | 1.21 (0.35) | 0.915 |
| C-reactive protein (mg/dL) | 0.68 (0.98) | 0.45 (0.68) | <0.001 | 0.59 (0.70) | 0.48 (0.76) | 0.023 |
| Uric acid (mg/dL) | 5.92 (1.57) | 5.37 (1.47) | <0.001 | 5.88 (1.55) | 5.39 (1.52) | <0.001 |
| **Liver chemistry** |  |  |  |  |  |  |
| Alanine aminotransferase (U/L) | 18.7 (12.2) | 23.4 (24.2) | <0.001 | 19.3 (13.4) | 23.5 (18.8) | <0.001 |
| Aspartate aminotransferase (U/L) | 22.6 (13.6) | 24.0 (23.2) | 0.065 | 23.1 (12.9) | 23.6 (12.4) | 0.577 |
| Gamma glutamyl transferase (U/L) | 41.4 (50.1) | 34.2 (39.5) | <0.001 | 49.6 (87.5) | 33.7 (29.8) | 0.001 |
| Alkaline phosphatase (U/L) | 97.7 (35.4) | 86.4 (25.0) | <0.001 | 97.5 (39.1) | 87.5 (31.0) | <0.001 |
| Total bilirubin (mg/dL) | 9.82 (5.24) | 9.85 (5.36) | 0.886 | 9.79 (4.64) | 9.96 (5.24) | 0.608 |
| Albumin (g/dL) | 40.3 (3.35) | 41.2 (3.43) | <0.001 | 40.2 (3.33) | 41.2 (3.45) | <0.001 |
| **Diabetes testing profile** |  |  |  |  |  |  |
| Glycated hemoglobin (%) | 6.29 (1.68) | 5.50 (1.11) | <0.001 | 6.25 (1.69) | 5.47 (1.03) | <0.001 |
| Fasting plasma glucose (mg/dL) | 123 (59.9) | 100 (36.7) | <0.001 | 124 (66.1) | 101 (38.1) | <0.001 |
| Fasting C-peptide (pmol/mL) | 6.81 (3.33) | 5.55 (2.04) | <0.001 | 6.87 (3.67) | 5.61 (2.12) | <0.001 |
| Fasting insulin (uU/mL) | 19.2 (23.7) | 15.4 (15.8) | <0.001 | 18.8 (19.0) | 15.3 (13.8) | 0.002 |
| **NAFLD indices** |  |  |  |  |  |  |
| NAFLD severity |  |  | <0.001 |  |  | 0.060 |
| Mild (%) | 278 (32.3%) | 573 (40.9%) |  | 126 (34.1%) | 250 (41.6%) |  |
| Moderate (%) | 370 (43.0%) | 559 (39.9%) |  | 150 (40.5%) | 221 (36.8%) |  |
| Severe (%) | 213 (24.7%) | 269 (19.2%) |  | 94 (25.4%) | 130 (21.6%) |  |
| NFS | -0.54 (1.51) | -2.01 (1.44) | <0.001 | -0.63 (1.48) | -2.14 (1.45) | <0.001 |
| FIB-4 | 1.32 (0.76) | 0.77 (0.46) | <0.001 | 1.30 (0.66) | 0.77 (0.81) | <0.001 |
| APRI | 0.25 (0.24) | 0.25 (0.30) | 0.893 | 0.25 (0.18) | 0.26 (0.46) | 0.650 |

NFS, NAFLD fibrosis score; FIB-4, Fibrosis-4 Score; APRI, aspartate aminotransferase-to-platelet ratio index.

### Model building and evaluation

Five ML methods, including LR, decision tree, RF, KNN and XGBoost, with all the 29 factors inputted were built for the mortality prediction of NAFLD patients. Absolute values of standardized beta coefficients for LR and feature importance for XGBoost and RF models were assessed and the results were shown in **Figure 2**. Age was the most important factor among all the ML models to predict mortality during follow-up period. SBP and glucose level were listed as the top 5 important variables of RF and XGBoost. The other essential factors for the models development were iron, transferrin saturation, BMI and uric acid for the LR, DBP and waist circumference for the RF, and HbA1c as well as C-peptide for the XGBoost.

**Figure 2.**
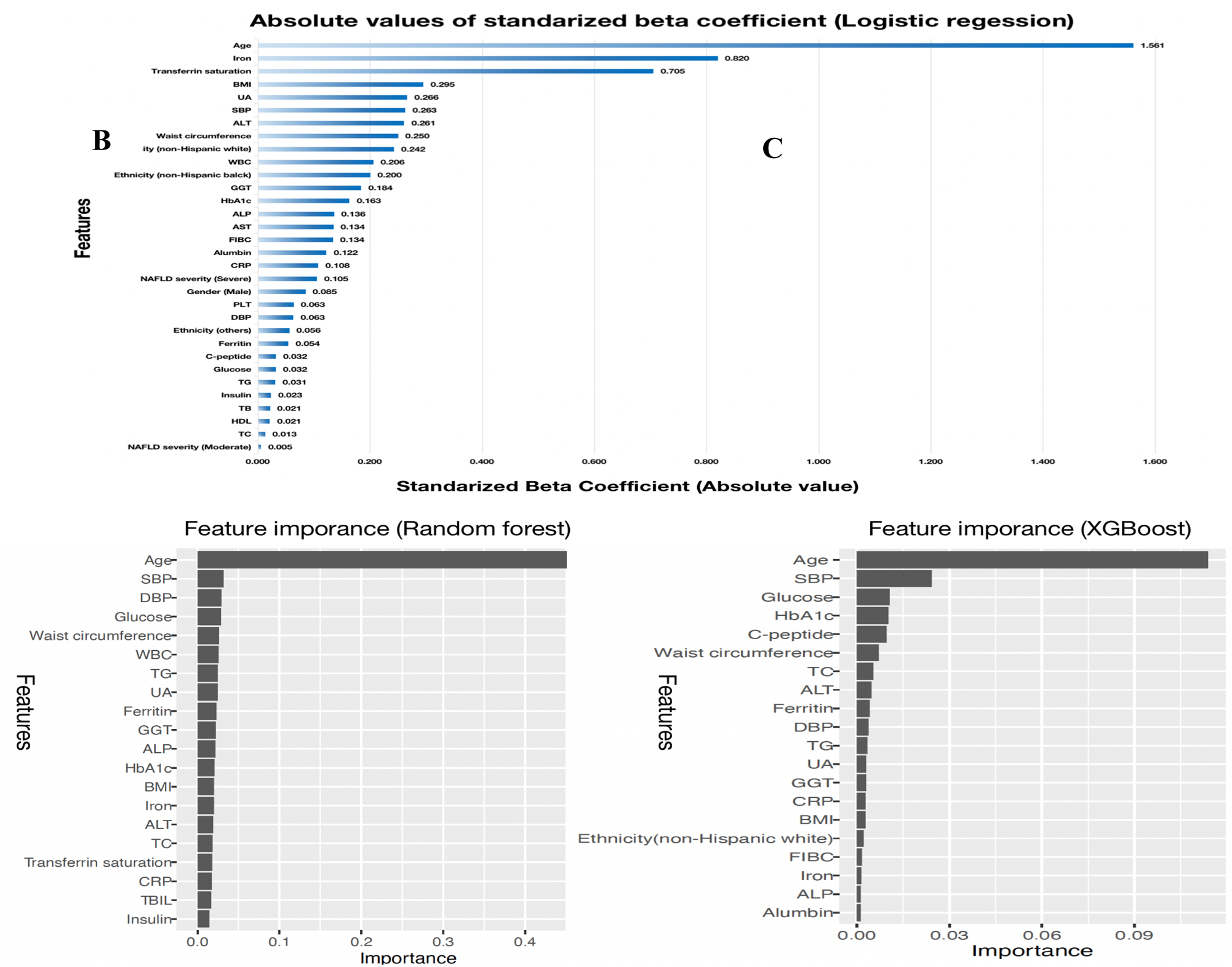
(A) Absolute values of standardized beta coefficients for the logistic regression model. (B) Feature importances of variables for the random forest model. (C) Feature importances of variables for the XGBoost model. BMI, Body mass index; UA, uric acid; SBP, systolic blood pressure; DBP, diastolic blood pressure; TC, total cholesterol; TG, triglyceride; HDL, high-density lipoprotein cholesterol; CRP, C-reactive protein; ALT, alanine aminotransferase; AST, aspartate aminotransferase; GGT, gamma glutamyl transferase; ALP, alkaline phosphatase; TBIL, total bilirubin; HbA1c, glycated hemoglobin; FIBC, total iron-binding capacity.

After 10-fold cross-validation, the training accuracy of ML models were 0.807 for the LR, 0.807 for the decision tree, 0.814 for the RF, 0.692 for the KNN, and 0.808 for the XGBoost.

### Models performance analysis

The receiver operating characteristic (ROC) curves with AUC values of the five ML models in the testing data is shown in **Figure 3**. Validation of our developed ML models showed reliable performance for the mortality prediction in NAFLD patients, whose AUC values were: LR, 0.888 (0.867–0.909); RF, 0.876 (0.852–0.897); XGBoost, 0875 (0.853–0.898); decision tree, 0.793 (0.766–0.819) and KNN, 0.787 (0.759–0.816), respectively. The F1 score of above models were 0.765, 0.759, 0.745, 0.744 and 0.759, respectively. The other evaluation measures of the prediction models, including accuracy, sensitivity, specificity, positive predictive value (PPV) and negative predictive value (NPV) are described in **Table 2**. Among all the evaluated classifiers, the LR model had the highest sensitivity of 0.819 and NPV of 0.878. The specificity, PPV and accuracy of the RF model were the highest on the other hand, which were 0.837, 0.745 and 0.813, respectively. **Table 2** shows the performance of NFS, FIB-4 and APRI on the testing data at the same time, and among all the conventional non-invasive scores, FIB-4 showed the best performance, whose accuracy was 73.0% and F1 score was 0.765. Nevertheless, the performance of all the ML models were superior to FIB-4 in all metrics.

**Figure 3.**
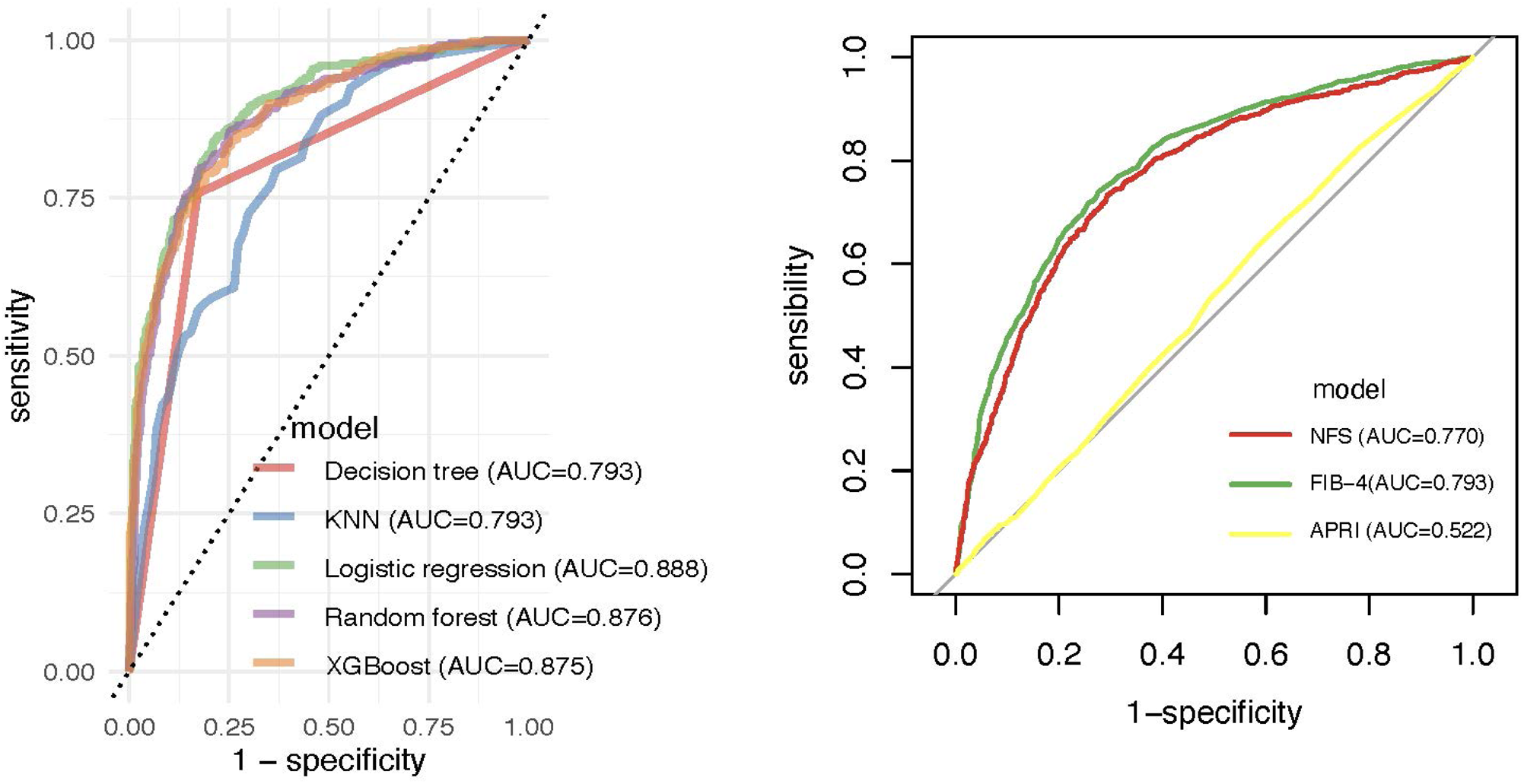
Comparison of ROC curves and AUC among the developed machine-learning models and among the conventional non-invasive scores for mortality prediction. ROC, receiver operating characteristic; AUC, area under the curve; NFS, NAFLD fibrosis score; FIB-4, fibrosis-4 score; APRI, aspartate aminotransferase-to-platelet ratio index.

**Table 2.** The performance of machine learning models and conventional non-invasive scores on testing data. AUC, area under the curve; CI, confidence interval; NPV, negative predictive value; PPV, positive predictive value; NFS, NAFLD fibrosis score; FIB-4, fibrosis-4 score; APRI, aspartate aminotransferase-to-platelet ratio index.

| Model | AUC [95%CI] | Accuracy (%) | PPV/Precision (%) | NPV (%) | Sensitivity/Recall (%) | Specificity (%) | F1 score |
| --- | --- | --- | --- | --- | --- | --- | --- |
| **Machine learning models** |  |  |  |  |  |  |  |
| Logistic regression | 0.888 [0.867-0.909] | 0.808 | 0.718 | 0.878 | 0.819 | 0.802 | 0.765 |
| Random forest | 0.876 [0.852-0.897] | 0.813 | 0.745 | 0.857 | 0.773 | 0.837 | 0.759 |
| XGBoost | 0.875 [0.853-0.898] | 0.803 | 0.736 | 0.846 | 0.754 | 0.834 | 0.745 |
| Decision tree | 0.793 [0.766-0.819] | 0.801 | 0.731 | 0.847 | 0.757 | 0.829 | 0.744 |
| KNN | 0.787 [0.759-0.816] | 0.813 | 0.745 | 0.857 | 0.608 | 0.737 | 0.759 |
| **Conventional non-invasive scores** |  |  |  |  |  |  |  |
| FIB-4 | 0.793 [0.777-0.809] | 0.730 | 0.622 | 0.819 | 0.739 | 0.724 | 0.675 |
| NFS | 0.770 [0.753-0.787] | 0.717 | 0.606 | 0.811 | 0.733 | 0.707 | 0.663 |
| APRI | 0.522 [0.502-0.543] | 0.499 | 0.400 | 0.649 | 0.634 | 0.416 | 0.491 |

Finally, **Supplementary Figure 1** shows the probability calibration curves of the ML models in validation and it can be seen that the predicted probability of all the models was uncertainty because they were not well-calibrated and underestimated.

## Discussion

To our knowledge, this study is the first time that the NAFLD mortality prediction model based on ML has been developed and evaluated. In conclusion, we selected 29 clinical variables for NAFLD mortality prediction from the NHANES-III database, which were important in the liver diseases and generally readily available at hospital admission. After using several ML algorithms of LR, decision tree, RF, KNN and XGBoost to train these variables, the verification of the developed models revealed dependable performance with relatively high AUC values. The LR model had both a higher AUC and F1 score, which indicated a superior performance in the death classification of NAFLD patients, and showed better performance than that of decision tree, RF, KNN and XGBoost.

On the other hand, we found that the decision tree model which consisted of only three factors: age, systolic blood pressure and HbA1c (**Figure 4**), although not showed the best performance among all the ML models, it had a certain degree of value (accuracy was 80.1%; AUC was 0.793 and F1 score was 0.744) in terms of testing performance. The decision tree model is easy to explain and use, so it can be used more practically in clinical practice.

**Figure 4.**
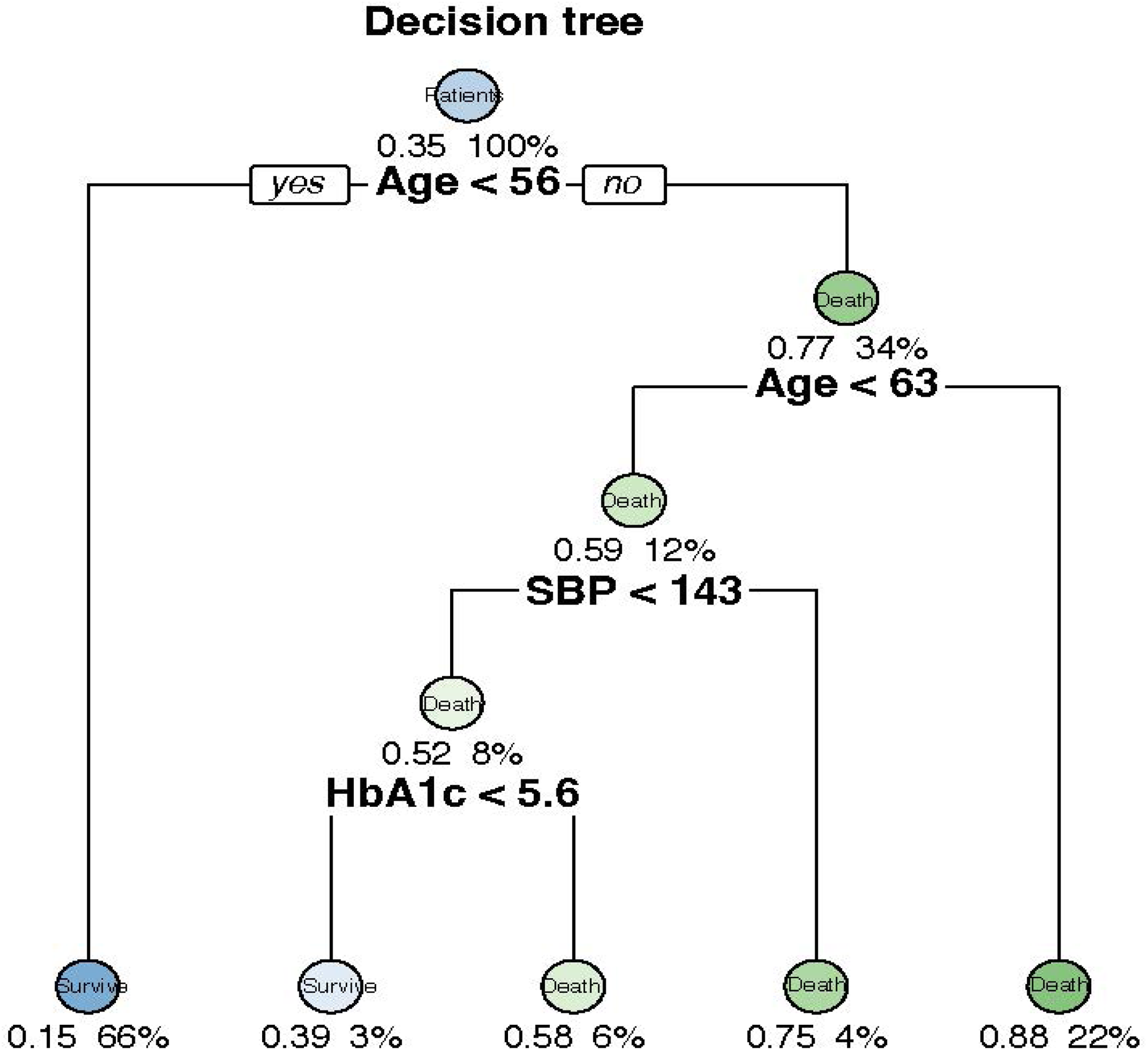
The decision logic of decision tree. SBP, systolic blood pressure; HbA1c, glycated hemoglobin.

The performance of some conventional non-invasive scores, like FIB-4, NFS and APRI for the overall mortality prediction in NAFLD patients was also showed in our study. AUC values for the overall mortality were: FIB-4, 0.793 (0.777–0.809); NFS, 0.770 (0.753–0.787) and APRI, 0.522 (0.502–0.543), respectively, which were closely similar to the results of a retrospective analysis of 646 biopsy-proven patients with NAFLD (AUC for the overall mortality were FIB-4, 0.72 (0.68–0.76); NFS, 0.72(0.68–0.76) and APRI, 0.52 (0.47–0.57)), indicating that these scoring systems were insufficient for clinical use.

On the other hand, the availability of ML methods in the development of medical prediction models have been proven in recent years^22, 23^. In the same way, by using a ML algorithm, a well-done prognostic model for NAFLD has been successfully developed in our study. With regard to the AUC values, the models we developed showed statistical advantages over the conventional non-invasive scores, and the F1 scores in the developed models were also significantly higher, which incited the validity of ML models in detecting NAFLD mortality. However, the calibration chart showed that the prediction of the probability of results was underestimated or overestimated, indicating that these models were only applicable to classification problems.

Our predictive models have the latent capacity for use in the clinical practice. Since we only used demographic characteristics and laboratory data as the predictor variables which are easily obtained, clinicians can use the predictive results as a reference tool to initiate treatment as early as possible. In addition, the model can be used to retrospectively evaluate the quality of care in NAFLD treatment. Nevertheless, ML models should not be used as an explicit tool to decide the withdraw of treatment.

Age is the most important factor not only in the decision tree model, but also ranks the first in all the other ML models. Many studies have proven that age is an independent risk factor for liver fibrosis in NAFLD patients^24, 25^, and liver fibrosis is also an independent risk factor for liver-related mortality^26–28^. Age is in correlation with increased cardiovascular mortality as the independent risk factor in NAFLD at the same time^29, 30^. So the role of age in all-cause mortality can be rationally explained.

Systolic blood pressure and HbA1c are also components of the decision tree to predict the all-cause mortality. With the deepening of research, people’s understanding of NAFLD is no longer limited to the liver itself, but as a major performance of metabolic syndrome (MetS) in the liver, which is closely associated with hypertension, obesity, dyslipidemia, type 2 diabetes (T2DM), insulin resistance (IR) and cardiovascular disease^31, 32^. Although patients with NAFLD, especially those with nonalcoholic steatohepatitis (NASH), have an increased risk of liver-related death, there is evidence that cardiovascular disease (CV) risk factors like hypertension, obesity, IR as well as T2DM are the major drivers of morbidity and mortality in NAFLD patients^33–35^. A recent study using NHANES-III database found poor glycaemic control (adjusted population-attributable fraction (PAF)=28.3% for all-cause mortality) and hypertension (adjusted PAF=23% for all-cause mortality) were the largest contributors to mortality for NAFLD patients and reaching desirable glycaemic control (HbA1c of <5.7%) could avoid 28.3% of all-cause deaths^36^.

Our study has several limitations. First, we all know that liver biopsy is the gold standard for NAFLD diagnosis, but it was ultrasound-proven in our study. Nevertheless, in population-based studies, it is the dominant imaging approach for NAFLD diagnosis and is available in primary care settings; Second, missing data is an unavoidable nature of NHANES III population dataset; Third, since we used the US registration database for training and verification of the model, we should use foreign databases for external verification in the future. However, there was no external data set like NHANES III that can be used to validate the model at the time of writing this paper; Fourth, due to the use of randomization in the modeling process, such as data segmentation, cross validation and the creation of some ML models, it may not be possible to completely reproduce the ML algorithm in our research. Finally, it might be criticized that ML models need a computing device to calculate results, and it is unrealistic to only use a single model for NAFLD patients. Since the features we selected are mainly patient background and laboratory data, we suggest that we can use ML models as a plugin for electrical health records after completing a prospective study of further performance improvement and future external validation.

But there are also some strengths in our study. First, to our knowledge, this study firstly assessed the performance of ML models in predicting all-cause mortality in NAFLD patients, based on over 3000 US individuals from NHANES III. Second, we proposed a simple model with rational performance of mortality prediction in patients with NAFLD, which will potentially be used by primary care providers in clinical practice.

## Conclusion

In conclusion, a new mortality prediction model for NAFLD patients in the USA was developed using ML technology. The LR model performed best in our study, using the AUC and F1 score for measurement. On the other hand, the decision tree model, which is composed of age, systolic blood pressure and HbA1c, can produce a rational prediction performance, and the most important thing is that it is the simplest to use. Although we need to further improve the quality of performance by increasing the sample size or conducting prospective validation in the clinical environment, our research has demonstrated for the first time the potential of NAFLD prediction models based on machine learning.

## Data Availability

ll relevant data are within the manuscript and its Supporting Information files.

## Author contributions

Study concept and design (BF), acquisition, analysis and interpretation of data, drafting of the manuscript (J-RZ), critical revision of the manuscript for important intellectual content (Z-LW), and study supervision (H-S C). All authors have made a significant contribution to this study and have approved the final manuscript.

